# Knowledge and perceptions of uterine fibroids: A descriptive cross-sectional survey among women of childbearing age in KwaZulu-Natal, South Africa

**DOI:** 10.1101/2024.03.18.24304504

**Authors:** Amanda Dlamini, Celenkosini Thembelenkosini Nxumalo, Nomakhosi Mpofana, Michael Paulse, Mokgadi Makgobole, Pavitra Pillay

## Abstract

**Background:** Uterine fibroids are the leading cause of hysterectomies among women of childbearing age. This study aims to elicit the knowledge, attitude and perceptions of childbearing women towards uterine fibroids in order to provide empirical evidence informing relevant interventions oriented toward health promotion in this regard.

**Methods:** A quantitative, cross-sectional descriptive design was used and data were collected from a sample of 362 women of reproductive age residing in a selected township in KwaZulu-Natal, South Africa. Ethical approval to conduct the study was obtained from the Durban University of Technology’s Institutional Research Ethics’ Committee (IREC – Ref No. BIREC 014/21). A pre-tested survey was conducted to gather data on knowledge, attitudes, and perceptions concerning uterine fibroids. The collected data were analyzed using SPSS version 27, employing descriptive statistics. Inferential statistics were also conducted to examine associations between key variables and respondents who self-reported being diagnosed with uterine fibroids.

**Results:** Most participants, 73.8% (n=267), had no awareness of uterine fibroids. Participants also demonstrated poor knowledge regarding the aetiology and symptoms of the condition. However, most participants, 49.2% (n=178), perceived uterine fibroids to be of spiritual origin, citing evil spirits and witchcraft as the cause. Participants subsequently reported that treatment would require herbal approaches and consultation with spiritualists such as traditional healers and seers. In summary, the study highlights various factors influencing self-reporting behaviours, including age, education level, employment status, marital status, number of children, awareness of the condition, perception of requiring treatment, family history, and symptom severity.

**Discussion and conclusion:** The study findings seem to suggest that women in the selected township lack accurate knowledge about uterine fibroids. These insights are valuable for shaping targeted health interventions and policies. Recognizing the complexities of self-reporting is crucial for improving health outcomes through early detection and tailored interventions.

## Introduction

Uterine fibroids are benign, smooth muscle growths of the uterus that are the most common forms of pelvic tumours among women (1). While the aetiology of fibroids remains relatively unknown, it is stated that the combination of hormonal and genetic factors are determinants of facilitative and synergistic factors that are influential in their growth (2, 3, 4). Although fibroids are often benign, they possess a combination of smooth muscle and fibroblastic components with a substantial presence of fibrous extracellular matrix which precipitates the pathogenic process (5). The location and size of fibroids in the uterus are important determinants of symptomatology and clinical manifestations (6). Nulliparity, race, obesity and familial history are common predisposing factors to clinically significant fibroids (2, 7, 8). The growth of uterine fibroids may be intramural, submucosal and sub-serosal (9). Uterine fibroids that are most common and often benign, grow intramurally, while the submucosal fibroids are least common and are often clinically significant (10).

Epidemiological data on uterine fibroids suggest that uterine fibroids occur in more than 70% of women at the onset of menopause (11, 12). Furthermore, research shows a higher incidence rate among Black women of African descent (2, 13). While the exact population incidence of uterine fibroids remains relatively unclear, there is substantial observational evidence indicating a higher incidence and prevalence rate among black women as compared to other races (14). The health outcomes and related complications owing to the occurrence of uterine fibroids is further exacerbated by socio-economic issues of level of education, employment and individual perceptual factors (15).

The clinical manifestations and symptoms of uterine fibroids vary, depending on the clinical significance. It is estimated that just under 50% of cases are often asymptomatic, (16) while the remaining cases often have symptoms ranging from mild to severe abdominal discomfort, acute pelvic pain, abnormal uterine bleeding, with variable amounts of bleeding depending on the size and location of the fibroid (17). Additional clinical features include frequency of micturition, swelling of the abdomen and increased pelvic pressure (18). Uterine fibroids may also result in complications leading to infertility, miscarriages, premature labour and complications related to prolonged uterine bleeding (19, 20). Child-bearing, pregnant women with uterine fibroids are also at higher risk of post-partum haemorrhage during labour and delivery (21, 22).

While treatment of fibroids depends on the size, location and nature of clinical symptoms (23, 24, 25), the use of oral contraceptives has been proven to be effective in providing some degree of protection against the development of uterine fibroids (26, 27, 28). The success of curative approaches for clinically significant fibroids depends on patient-related and health service-related factors among the many determinants. In the instances of benign asymptomatic factors, patient and health system factors may also be said to be influential in determining the course and health outcomes related to the presence of such types of fibroids. In both of the aforementioned instances, the patient as the key determining factor cannot be understated. The notion of the patient as a critical determining factor relates to an accurate, comprehensive and holistic understanding and awareness regarding uterine fibroids (29). This means that awareness, knowledge, attitudes and perceptions extend beyond mere recognition of signs and symptoms but also encompass an understanding of the nature and origins of fibroids, coupled with appropriate behaviour change interventions that relate to prevention and health seeking behaviour in order to attain relevant treatment as necessitated by clinical significance.

Since reproductive and general health outcomes related to the presence of fibroids are dependent on individual patient factors of knowledge, attitudes and perceptions, an awareness of these factors is essential among women of reproductive age so that uptake of relevant health seeking interventions may be facilitated in order to initiate appropriate treatment and reverse potential complications. While the knowledge, awareness and perceptions of women regarding uterine fibroids has been assessed and explored globally (30, 31, 32), there is a paucity of such data among women of reproductive age, particularly in sub-Saharan Africa, more specifically, in South Africa. Moreover, there is a dearth of context specific research, meaning that contextual and evidence-informed education interventions related to fibroids remain suboptimal for women of reproductive age. The aim of this study was thus to assess the knowledge, attitudes and perceptions towards uterine fibroids among women of childbearing age in rural KwaZulu-Natal, South Africa, to facilitate contextual awareness regarding existing knowledge, attitudes and perceptions concerning uterine fibroids. These findings subsequently have implications for informing educational interventions to facilitate awareness regarding uterine fibroids. Ghant et al (2016)(31) argue that there is a need for patient-centred and community-based interventions to improve women’s knowledge regarding uterine fibroids so that relevant treatment options may be promoted. It is postulated that awareness may result in positive health-seeking behaviour related to prevention and treatment of fibroids. This may subsequently have positive outcomes on quality of life and public health, because fibroids are the leading cause of high rates of total hysterectomies among childbearing women (33).

## Materials and Methods

This study employed a quantitative, cross-sectional descriptive design to assess the knowledge, attitudes and perceptions of childbearing women aged 18 to 35 towards uterine fibroids in a township in KwaZulu-Natal, South Africa. Data were collected between 19 July to 30 October 2021. Non-probability convenience sampling was used to recruit participants for the study. Three hundred and sixty-two (*N* = 362) childbearing women were enrolled in the study based on the sample size calculated using the G*Power software version 3.1.9.7 (34).

This is line with suggestions made by various statistical scholars: Cohen (1992) suggests that a priori sample size determination is necessary for sufficient power in chi-squared tests and statistical power analysis is essential to guarantee that studies can detect effects (35). The non-centrality parameter affects these tests’ power and needed sample size, reflecting the expected test statistic in the event of an alternate hypothesis (36). Standard large-sample formulas can approximate power, sample size and the smallest detectable effect (37); conventional effect size values and their corresponding sample sizes for achieving a power of 0.80 offer guidelines (38). To prevent underpowered studies, effect size estimates from prior research should take uncertainty into account (39). Considering all of this, G*Power yielded a minimal acceptable size of 32 to 55 for degrees of freedom, *df*, ranging from 1 to 6 respectively for a large effect size, *w* = 0.50, α err prob = 0.05, power (1-β) err prob = 0.80 which is substantially lower than what was finally sampled.

Childbearing women were recruited to participate in the study at community level through using random selection, based on convenience sampling approaches applied at the primary researcher’s discretion. A structured, pre-tested interviewer-administered questionnaire was used to collect data from participants. The structured questionnaire comprised questions to elicit demographic details of participants such as age, level of education, occupation and marital status. Specific questions of knowledge and awareness related to signs, symptoms, risk factors and complications of uterine fibroids were also detailed in the questionnaire.

The data collected were entered on to Microsoft Excel and analysed using Statistical Product and Service Solutions version (SPSS), version 27. Initially, exploratory descriptive statistics were conducted to analyze the frequencies of various categories, behaviours, attitudes, and medical conditions among participants. This step provided a comprehensive overview of the distribution of variables in the dataset, allowing researchers to identify patterns and trends.

Following the exploratory descriptive analysis, chi-squared tests were employed to investigate associations between these variables and participants’ self-reporting of a diagnosis of uterine fibroids. chi-squared tests are commonly used for analyzing categorical data and determining whether there is a significant association between variables (40). They assess whether the observed frequencies in different categories deviate significantly from what would be expected by chance (41). In this study, chi-squared tests were chosen due to their ability to handle categorical data and their non-parametric nature, making them suitable for analyzing variables with non-normal distributions (42, 43). The results of the chi-squared tests provided insights into the relationships between various factors and participants’ likelihood of self-reporting a diagnosis of uterine fibroids.

However, it’s important to note that while chi-squared tests are widely used, they have limitations, particularly in cases where sample sizes are small or when the assumptions of the test are violated (44, 45, 46). In instances where the expected frequencies are low, the Fisher’s exact test may be more appropriate as it provides more accurate results (47, 48, 49). Despite this, the initial exploratory descriptive statistics provided valuable context for the subsequent chi-squared analyses, enhancing the overall understanding of the relationships between variables in the study.

### Ethical Approval and Considerations

The study was conducted in accordance with the Declaration of Helsinki, and approved by the Durban University of Technology’s Institutional Research Ethics’ Committee (BIREC 014/21). A written informed consent was obtained from all participants prior to data collection and all ethical principles of research were observed when conducting the study.

## Results

### Participant Demographics

Table 1 presents the demographic characteristics of the participants involved in the study on knowledge and perceptions of uterine fibroids among women of childbearing age in KwaZulu-Natal, South Africa.

**Table 1.** Participant Demographics.

| Characteristics | Frequency<br>(N) | Percentage<br>(%) |
| --- | --- | --- |
| ***Age*** |  |  |
| 18-25 | 227 | 62.7 |
| 26-31 | 30 | 8.3 |
| 32-40 | 105 | 29 |
| ***Level of Education*** |  |  |
| Illiterate | 8 | 2.2 |
| Primary School Education | 6 | 1.7 |
| Read and write | 5 | 1.4 |
| Secondary School Education | 69 | 19.1 |
| Tertiary Education | 274 | 75.7 |
| ***Marital status*** |  |  |
| Divorced | 16 | 4.4 |
| Married | 69 | 19.1 |
| Single | 254 | 70.2 |
| Widowed | 23 | 6.4 |
| ***Occupation*** |  |  |
| Unemployed | 295 | 81.5 |
| Employed | 67 | 18.5 |
| TOTAL | 362 | 100 |

### Age

Participants were categorized into three age groups: 18-25, 26-31, and 32-40 years. The majority of participants fell within the age range of 18-25 years (62.7%), followed by 32-40 years (29.0%), and 26-31 years (8.3%).

### Level of Education

Participants’ educational attainment varied, with the majority having tertiary education (75.7%). A smaller proportion had secondary education (19.1%), while a very small percentage had either primary education (1.7%) or could only read and write (1.4%). A minority reported being illiterate (2.2%).

### Marital Status

Participants’ marital status was diverse, with the majority being single (70.2%). Married women made up (19.1%) of the sample, while smaller percentages were either widowed (6.4%) or divorced (4.4%).

### Occupation

The majority of participants reported being unemployed (81.5%), while a smaller proportion were employed (18.5%).

These demographic characteristics provide insight into the composition of the participant sample in terms of age, education, marital status, and occupation, which is crucial for understanding the population under study and interpreting the findings of the research accurately

In addition, the study explored associations between participant demographics, perceptions, lifestyles, and self-reported diagnosis of uterine fibroids were examined. It is important to note that associations do not necessarily imply causation but allow for future research exploration (40, 50, 51, 52). Table 2 displays the association between various categories of interest and participants’ self-reported diagnosis of uterine fibroids, along with the chi-square statistics for each category. The analysis reflects the following:

**Table 2.**
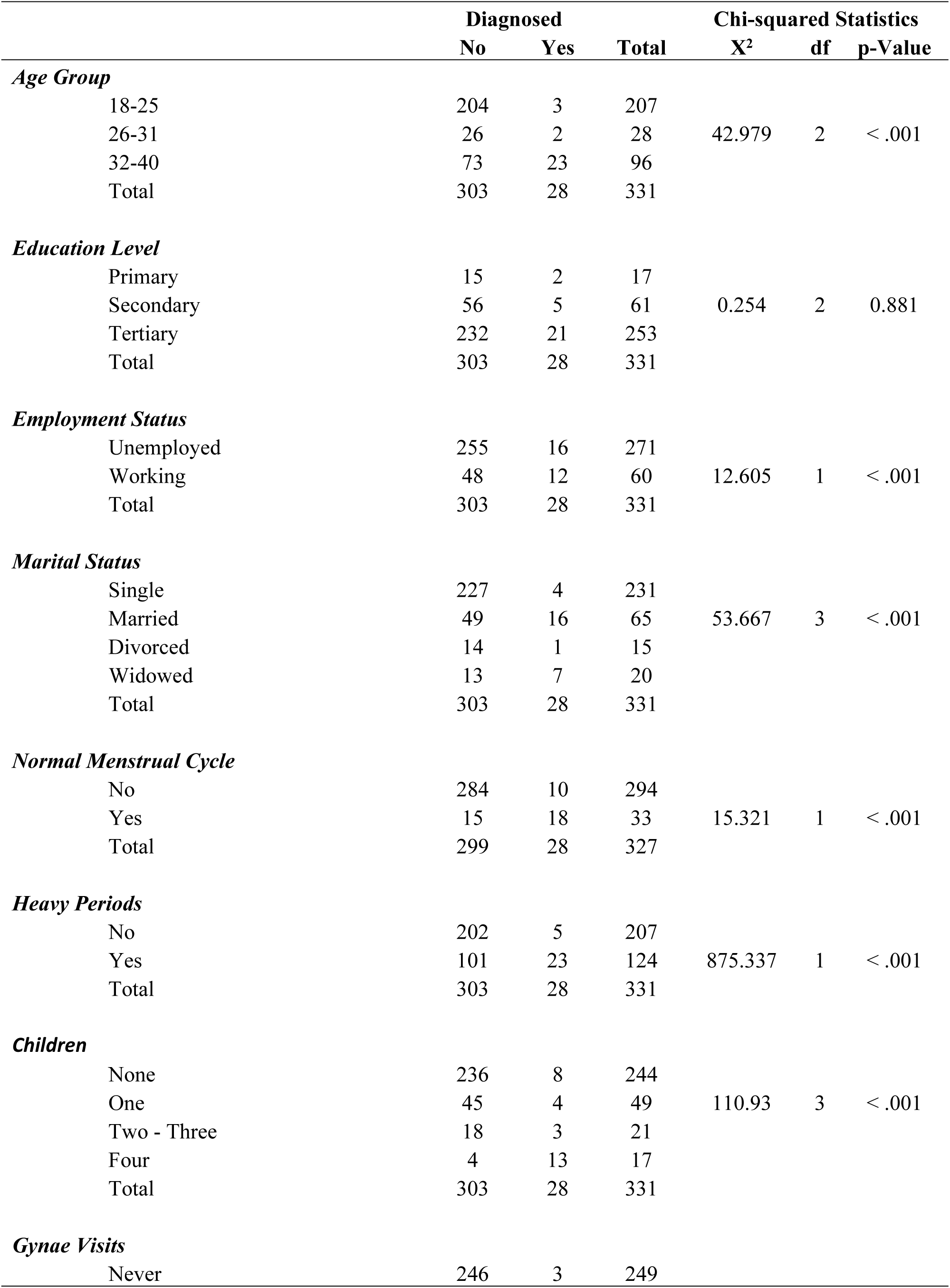
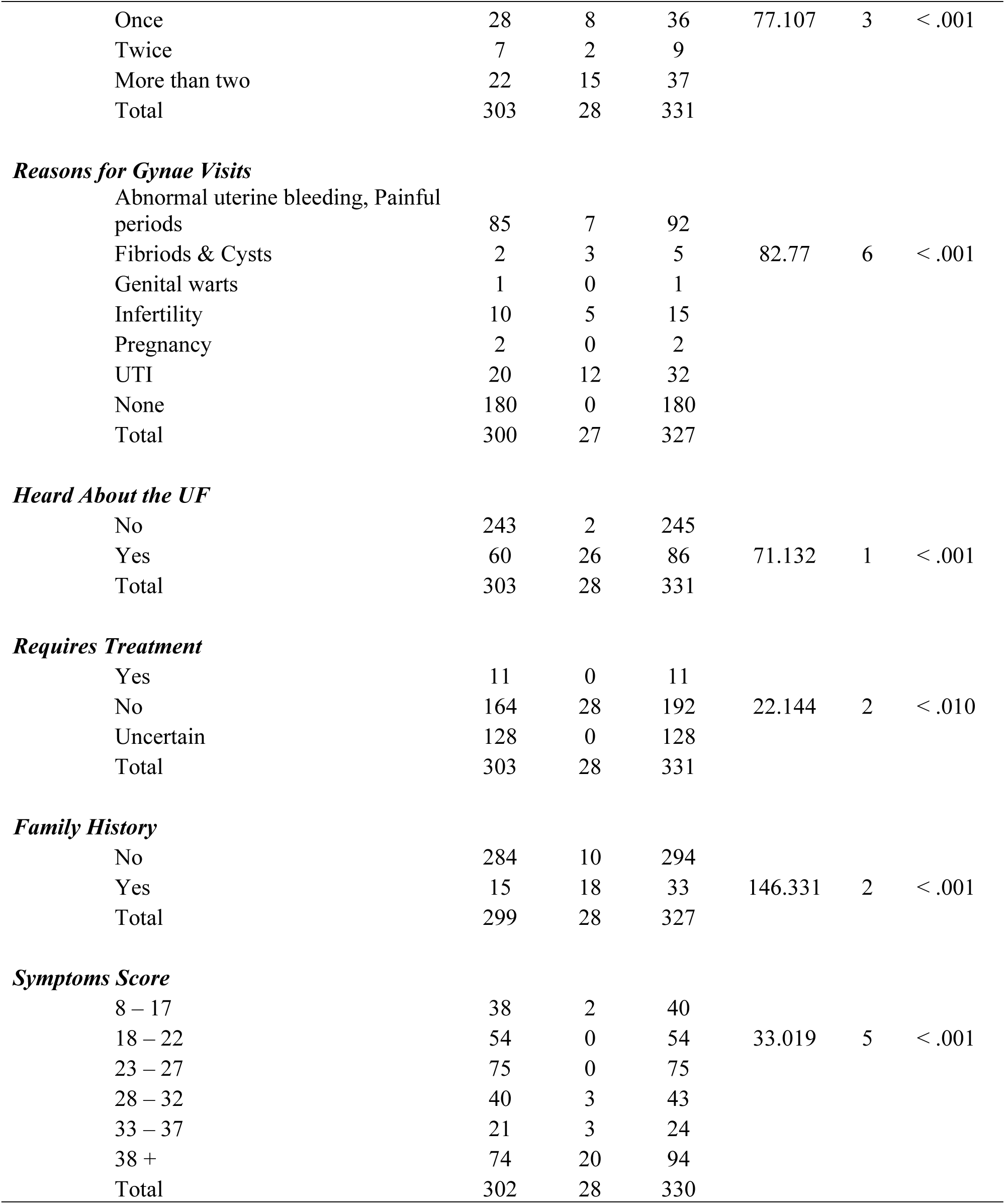
Association of Categories of Interest and the Participants’ Self-Reported Diagnosis.

### Age Group

A significant association was found between age group and self-reported diagnosis of uterine fibroids (χ2 = 42.979, df = 2, p < .001). Specifically, within the 26-31 age group, there were significantly more self-reported diagnoses compared to the other age groups.

### Education Level

Participants’ education levels were classified as primary, secondary, or tertiary. The analysis revealed no significant association between education level and self-reported diagnosis of uterine fibroids (χ2 = 0.254, df = 2, p = 0.881).

### Employment Status

Participants were categorized as unemployed or working. Employment status showed a significant association with self-reported diagnosis of uterine fibroids (χ2 = 12.605, df = 1, p < .001). A higher proportion of working participants reported a diagnosis compared to unemployed participants.

### Marital Status

Marital status was classified as single, married, divorced, or widowed. A significant association was found between marital status and self-reported diagnosis of uterine fibroids (χ2 = 53.667, df = 3, p < .001). Specifically, married participants were more likely to report a diagnosis compared to participants in other marital status categories.

### Normal Menstrual Cycle

Participants were asked about the regularity of their menstrual cycles. There was a significant association between having a normal menstrual cycle and self-reported diagnosis of uterine fibroids (χ2 = 15.321, df = 1, p < .001). Participants with a normal menstrual cycle were more likely to report a diagnosis.

### Heavy Periods

Participants were asked whether they experienced heavy periods. Heavy periods showed a significant association with self-reported diagnosis of uterine fibroids (χ2 = 875.337, df = 1, p < .001). Participants experiencing heavy periods were overwhelmingly more likely to report a diagnosis.

### Children

The number of children participants had was categorized as none, one, two to three, or four. There was a significant association between the number of children and self-reported diagnosis of uterine fibroids (χ2 = 110.930, df = 3, p < .001). Participants with four children reported the highest proportion of diagnoses.

### Gynaecological Visits

Participants’ frequency of gynaecological visits was categorized as never, once, twice, or more than twice. A significant association was found between the frequency of gynaecological visits and self-reported diagnosis of uterine fibroids (χ2 = 77.107, df = 3, p < .001). Participants who visited a gynaecologist more than twice were more likely to report a diagnosis.

### Reasons for Gynaecological Visits

Various reasons for gynaecological visits were explored. A significant association was found between the reason for visits and self-reported diagnosis of uterine fibroids (χ2 = 82.770, df = 6, p < .001). Particularly, visits due to abnormal uterine bleeding and painful periods were strongly associated with reporting a diagnosis.

### Awareness of Uterine Fibroids

Participants were asked if they had heard about uterine fibroids. There was a significant association between awareness of uterine fibroids and self-reported diagnosis (χ2 = 71.132, df = 1, p < .001). Participants who had heard about uterine fibroids were more likely to report a diagnosis.

### Perceived Need for Treatment

Participants were asked if they perceived a need for treatment for uterine fibroids. There was a significant association between perceived need for treatment and self-reported diagnosis (χ2 = 22.144, df = 2, p < .010). Participants who reported a diagnosis were more likely to perceive a need for treatment.

### Family History

Participants were asked about a family history of uterine fibroids. A significant association was found between family history and self-reported diagnosis (χ2 = 146.331, df = 2, p < .001). Participants with a family history of uterine fibroids were more likely to report a diagnosis.

### Symptoms Score

Participants’ symptom scores were categorized into six groups. There was a significant association between symptom score and self-reported diagnosis (χ2 = 33.019, df = 5, p < .001). Participants with higher symptom scores were more likely to report a diagnosis of uterine fibroids.

These findings highlight the importance of demographic factors, health behaviours, and symptomatology in the diagnosis and perception of uterine fibroids among women of childbearing age in KwaZulu-Natal, South Africa.

### Knowledge of uterine fibroids

To elicit participants’ knowledge about uterine fibroids, open-and closed-ended questions were posed concerning awareness, causes and definition of uterine fibroids. Table 3 shows that regarding awareness of uterine fibroids, most participants, 76% (n=276), indicated that they had never heard of uterine fibroids.

**Table 3.**
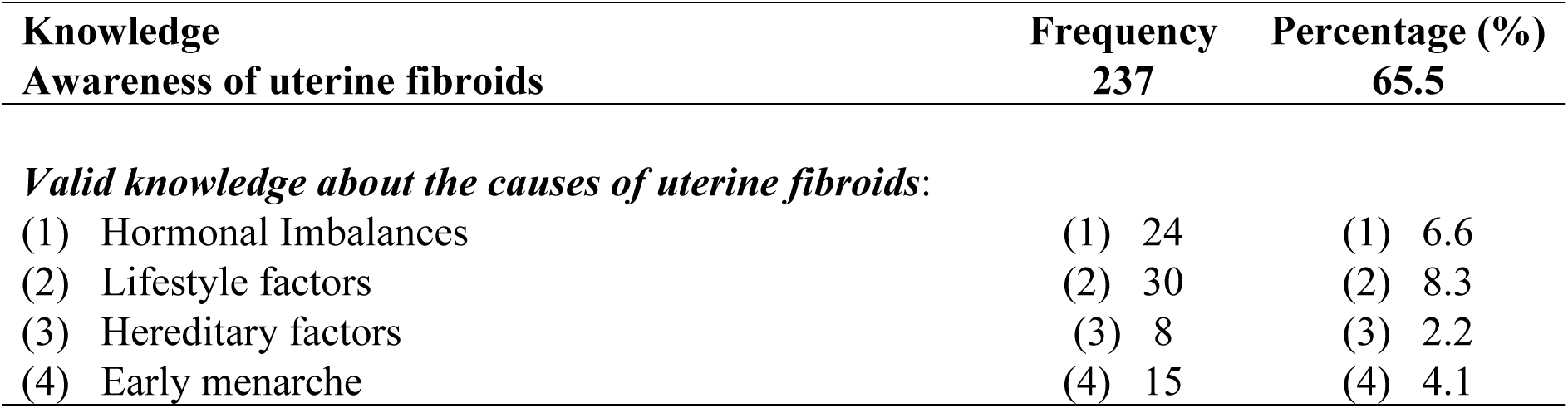
Knowledge of Uterine Fibroids.

This finding was subsequently confirmed by the results of the binominal test which was conducted as a follow-up, highlighting that this finding was statistically significant. When asked about the causes of uterine fibroids, 65. 5% (n=237) did not provide a response, while an additional 7.7% (n= 28) provided an invalid response. The remaining participants provided valid responses, citing various answers to this question, as 6.6% (n=24) revealed that uterine fibroids are caused by hormonal imbalances; 8.3 % (n=30) cited lifestyle factors such as obesity, high stress levels and multiple sexual partners as a cause. A further 2.2% (n=8) cited hereditary factors, while 4.1% (n=15) stated that uterine fibroids result from the onset of early menarche. Another 0.8% (n=3) of the participants stated that uterine fibroids are caused by evil spirits. To ascertain knowledge of uterine fibroids by definition, participants were asked an open-ended question which yielded responses that were categorised into different subgroups. Only 14,9% of the participants (n=54) responded to the aforementioned question and defined uterine fibroids as a type of cancer.

### Perceptions of uterine fibroids

To elicit responses of perceptions related to uterine fibroids, participants were asked questions related to the perceived age group that is normally affected; the treatment approaches that are most effective or suitable, and perceptions regarding the origin of uterine fibroids. Table 4 highlights participants responses regarding the perceived age group that is normally affected by uterine fibroids, 24.9%(n=90) indicated that uterine fibroids occur in females of ten (10) years and older, whereas 8.3% (n=30) revealed that uterine fibroids affect any age group. The remaining participants, 62,7% (n=227), did not provide a response to this question.

**Table 4.** Perceptions of uterine fibroids.

| Perception | Frequency | Percentage (%) |
| --- | --- | --- |
| Uterine fibroids affect females >10 years | 90 | 24.9 |
| Uterine fibroids affect females in any age group | 30 | 8.3 |
| Uterine fibroids require treatment interventions | 200 | 55 |
| Uterine fibroids require spiritual treatment | 178 | 49.2 |
| Uterine fibroids require medical treatment | 31 | 8.6 |
| Uterine fibroids require traditional and/or herbal treatment | 153 | 42.3 |

About 55% of the participants (n=200) agreed that uterine fibroids require treatment and 3.3% (n=12) stated that they did not require treatment. Of the participants, 49.2% (n=178) indicated that spiritual treatment (prayer) is required to manage uterine fibroids, while 42.3% (n=153) stated that herbal remedies were required to cure uterine fibroids. Only 8.6% (n=31) of the participants believed that medical treatment was required.

## Discussion

This study assessed the knowledge, attitudes and perceptions of childbearing women regarding uterine fibroids in KwaZulu-Natal, South Africa. The study findings revealed that the majority of participants in this study had never heard of uterine fibroids, implying a lack of adequate knowledge among these participants. This finding concurs with previous studies (53, 54) where low levels of knowledge were also observed among women, including those with reasonably acceptable levels of education. A lack of knowledge concerning uterine fibroids was noted, since the majority of participants indicated that these are cancerous growths and painful periods, while few participants displayed an awareness that these are non-cancerous. The reported misconceptions corroborate the study conducted by Saghir, Kamran (55) which indicated that participants incorrectly define uterine fibroids (ufs). In this study, the majority of participants perceived the common age occurrence of ufs to be in the range of 10 years and older and in any age group, which was not the expected response. This finding contradicts many studies, including that of Borah, Yao (56) which indicated the common age occurrence as 18-54. However, Gao and Wang (57) suggested that the common age occurrence is over 35 years .

Although the aetiology of uterine fibroids remained elusive, there was an indication of knowledge of the predisposing factors to uterine fibroids (58). The majority of the participants showed a correct knowledge of the predisposing factors, which included stress, obesity, hereditary, early menarche, caffeine, smoking and hormones. This is consistent with the studies of (59, 60, 61). A few participants indicated that uterine fibroids are caused by having multiple partners, although a study conducted by Azzahra, Gondodiputro (62) proved that there is no association between this factor and uterine fibroids. It was also noted that participants indicated that uterine fibroids are caused by evil spirits. This corroborates findings in the study conducted by Akpenpuun *et al.* (2019).

Most participants indicated that uterine fibroids require treatment, while a few said they do not require treatment. The study conducted by Ciebiera, Ali (64) also indicated similar findings. Around two-fifths (2/5) of the participants indicated that uterine fibroids are a spiritual problem and that prayer is used in treating this disease. The study conducted by Wu, Shao (65) also indicated similar findings. However, these findings contradict the findings of Arisukwu, Nwogu (66) in which celibate-women, despite their spiritual lifestyle, did not embrace praying and fasting as options for fibroid prevention. A further two-fifths (2/5) of the participants indicated that herbal treatment is required for the treatment of uterine fibroids. This is consistent with the observations of Arisukwu, Nwogu (66) that, regardless of their level of education, women still opt for local herbs as a form of treatment. The remaining one-fifth (1/5) of the participants indicated that medical treatment is ideal for uterine fibroids which concurs with the findings of Millien, Manzi (67).

A study conducted by Ikechebelu, Okpala (68) indicated that women with uterine fibroids frequently experience multiple gynaecological pain symptoms and bleeding symptoms, compared to women with no uterine fibroids. Riggan, Stewart (69) stated that most women lack knowledge of uterine fibroids symptoms. The strong correlation across the symptoms indicates that participants are likely to experience all the symptoms. These findings are consistent with those observed in the study conducted by Tojieva, Khalimova (60) and that of Singh, Shinde (70).

It was observed during the analysis, that the majority of the participants in the present study indicated that they experienced heavy bleeding during menstrual periods. A study conducted by Naz, Memon (71) indicated that women with uterine fibroids are two (2) to three (3) times more likely to experience heavy bleeding during menstrual periods than those who do not have uterine fibroids. These findings corroborate the study conducted by Fuldeore and Soliman (18). Anaemia, which is a possible consequence of severe menstrual bleeding, was moderate among the participants. This was expected, hence heavy menstrual bleeding is one of the hallmark symptoms of uterine fibroids (69).

Most participants in this study indicated that they experienced the passing of blood clots during menstrual periods. This is consistent with the study conducted by Swain, Yadav (72) where blood clots were reported by women with uterine fibroids. These findings suggest an association between blood clot passing and uterine fibroids which is supported by the study of Salas, López (73).

Fluctuation in the duration of menstrual periods compared to previous cycles was reported by the most of the participants in this study and is associated with uterine fibroids. Many studies are in agreement with this observation that women with uterine fibroids experience fluctuation in the duration of menstrual periods compared to previous cycles (74, 75). The majority of the participants indicated that they experienced fluctuation in the length of monthly cycles compared to previous cycles which is associated with uterine fibroids. This corroborates the study conducted by Tayebi, Izaddost (74) which also observed fluctuation in the length of monthly cycles compared to previous cycles in women with uterine fibroids. More than two-thirds (2/3) of the participants indicated that they experienced pressure in the pelvic area. This could be the reason why the majority of participants reported sexual dyspareunia (pain during sexual intercourse) since this is a complication caused by pressure in the pelvic area (76). These findings are consistent with the findings observed by Xie *et al.* (2020) that women with uterine fibroids reported pelvic pressure.

The present study findings imply that most of the participants experience urinary problems. This suggests that these problems may be caused by uterine fibroids. The study by Laughlin-Tommaso, Lu (77) also observed similar findings where participants experienced frequent urination. Participants reported that they experienced fatigue; which is a symptom associated with uterine fibroids. Fatigue could also be associated with emotional problems which were reported by some of the participants (78). These findings corroborate those observed in the study conducted by Harmon (79).

Infertility was reported by a few participants in this study. It is believed that fibroids can result in infertility due to their location and size (80). There are many studies which indicate that uterine fibroids are mostly seen in nulliparous women (62, 81), however, it was observed in a study conducted by Saghir, Kamran (55) that multiparous women also experience uterine fibroids due to early pregnancy. A few participants reported abortion as a problem, which is suggestive that this problem might be caused by uterine fibroids. Tran, Al Naber (82) indicated that that women with uterine fibroids may experience spontaneous abortion according to the size and location of the fibroids. These findings corroborate those observed in the study conducted by Fortin, Flyckt (83).

### Study Limitations

The study design only included questionnaire data on the knowledge and perceptions of uterine fibroids, which could not be confirmed with patient medical records. The study was aimed at women of childbearing age only and did not include the views of medical staff that may be responsible for managing or treating uterine fibroids. The study also did not consider the available health care facilities for women at risk to approach should they experience symptoms of complications related to uterine fibroids.

## Conclusion

The aim of this study was to assess the knowledge and perceptions regarding uterine fibroids among women of childbearing age in KwaZulu-Natal, South Africa, to provide contextual awareness regarding existing knowledge and perceptions on this subject matter. The findings suggest that women of 18-40 years of age in this study lack knowledge of uterine fibroids, hence they live chronically with symptoms, without seeking help. It appears that limited knowledge of uterine fibroids and normal menstruation can lead to a distorted view of what constitutes normality concerning uterine bleeding. The findings imply that further studies to investigate individualised and community-based education interventions related to uterine fibroids are necessary to promote self-preventive interventions, as well as health-seeking behaviours among women. While there seems to be a more profound focus on other female health issues such as cervical cancer and breast cancer, knowledge and awareness of uterine fibroids among communities at risk are also needed as this could help alleviate the related complications among women of childbearing age in KwaZulu-Natal.

## Data Availability

Due to the sensitive nature of the data, it will be kept by the researchers and made available on demand.

## Acknowledgments

We would like to thank all the participants for their enthusiastic participation in this study.

## Data availability

Due to privacy, data are available upon request from the first author.

